## Supplementary Information for "Non-linear genetic regulation of the blood plasma proteome"

**Supplementary Note 1: Variant pre-filtering using GWAS**

We implemented variant pre-filtering by GWAS into the EIR-auto-GP pipeline to reduce the number of input variants in the DL model to prevent overfitting and reduce computational costs (**Methods**). For each protein, a GWAS using PLINK2[27] was performed to identify pQTL associations. The results of the GWAS were overall concordant with results from Sun et al. despite different input variants (**Figure 1b**). Specifically, while Sun et al., identified 14,287 significant (P<1.7e-11) associations when using 16.1 million variants from imputed data, we identified 172,854 significant (P<1.7e-11) associations among 49,276 unique variants when using 424,097 autosomal variants from called genotypes (**Supplementary Data 1**). We found their distribution across the genome closely matched that of the previous study. For example, pQTLs on chromosomes 6, 9, 10, and 19 were highly associated with protein levels across the whole proteome and overlapping SNVs highly correlated with cor=0.96, pval=2e-308 (**Supplementary Figure 1b, Figure 1b**). The discrepancy in the number of significant hits was likely due to analyzing called genotypes, not applying LD pruning, and that we used a different tool for the GWAS, i.e., PLINK2[27] as opposed to REGENIE[28]. The choice of PLINK2, which employs a simple linear regression approach, could contribute to a higher proportion of false positives compared to REGENIE. REGENIE approximates a mixed model, which is generally more effective at controlling for false positives. Most (67.2%) variants were only associated with a single protein, with some, such as the ABO intron variant rs507666, associating with 249 different protein levels (**Supplementary Figure 1c**). We found most proteins to be associated with fewer than 100 variants, with HLA-A being the protein with the highest number of associated variants (3,624) (**Supplementary Figure 1d**).

**Supplementary Note 2: High modeling performance was associated with concordant Olink and SomaScan measurements**

Multiple studies have compared antibody (Olink) and aptamer-based (SomaScan) proteomic profiling methods and found discrepancies in some protein measurements[4,37]. We, therefore, compared the correlation coefficients of Olink and SomaScan for 1,861 proteins from Eldjarn et al., with the performance of our linear and DL models. We found that proteins with low performance in our models ($R^2<0.1$) had a low correlation between Olink and SomaScan (Wilcoxon rank-sum test, linear: W=326121, p=5.77e-14; DL: W=325089, p=1.511e-14) (**Supplementary Figure 2g**). For example, of the proteins with $R^2<0.1$ using our DL model, 406 (21.8%) showed a Spearman correlation between Olink and SomaScan of <0.1, whereas only 182 (9.8%) of proteins with DL $R^2 \geq 0.1$ showed a Spearman correlation of <0.1 (**Supplementary Figure 2h**). This indicated that the low performance of some proteins in the DL and the linear models could be due to noisy protein measurements. Similarly, we investigated whether the type of Olink panel (Oncology, Neurology, Inflammation, Cardiometabolic) influenced the comparison between the DL and linear model. Here, we did not observe differences in performance gap (DL-linear) between the proteins from different Olink panels (**Supplementary Figure 2i**). However, we found that proteins from the first panels performed better using both DL and linear models than proteins from the second panels (t-test, two-sided, t = 17.109, df = 5739.9, p-value < 2.2e-16) (**Supplementary Figure 2j**). This was in line with results from Sun et al., who found a decreasing number of new associations with an increasing number of proteins measured[3].

### Supplementary Figure 1

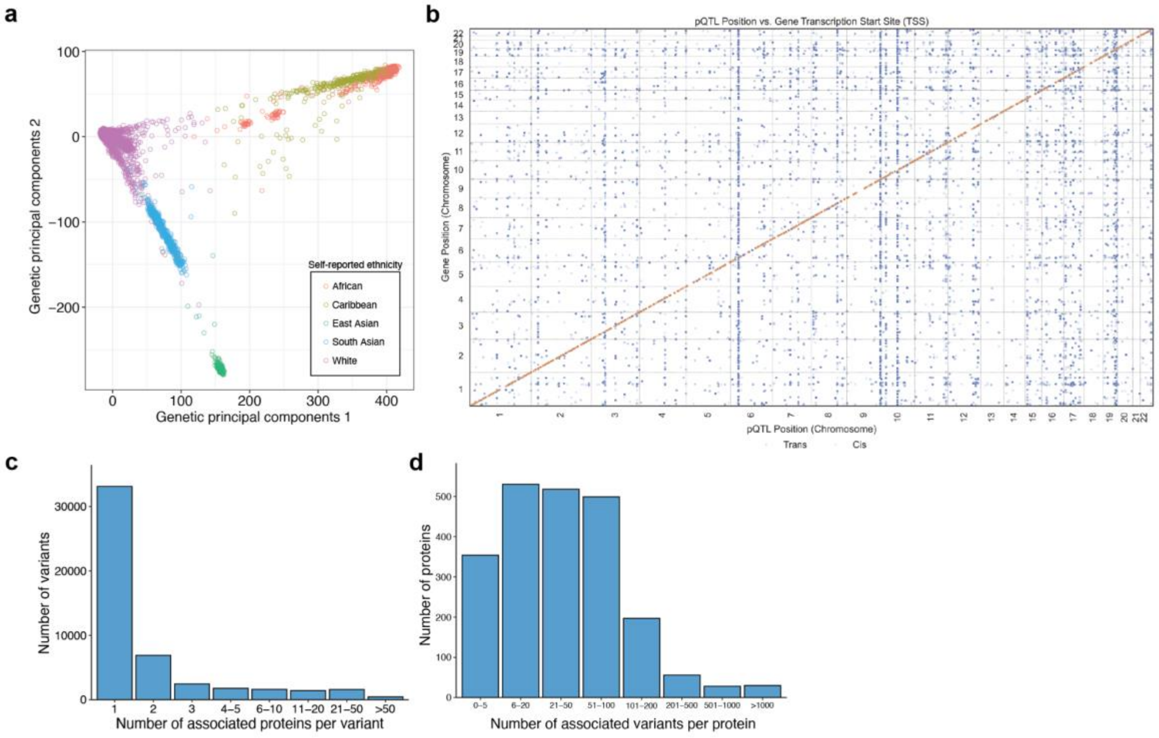


**Supplementary Figure 1. a)** Genetic Principal Components for 52,700 participants in the UKB-PPP stratified by grouped self-reported ethnicities. **b)** pQTL and gene position of significant (p<5e-11) pQTL identified by GWAS for 2922 proteins. Red indicates cis-pQTL and blue indicates trans-pQTL. **c)** Number of significantly associated proteins per variant identified by GWAS. **d)** Number of significant associations per protein identified by GWAS.

### Supplementary Figure 2

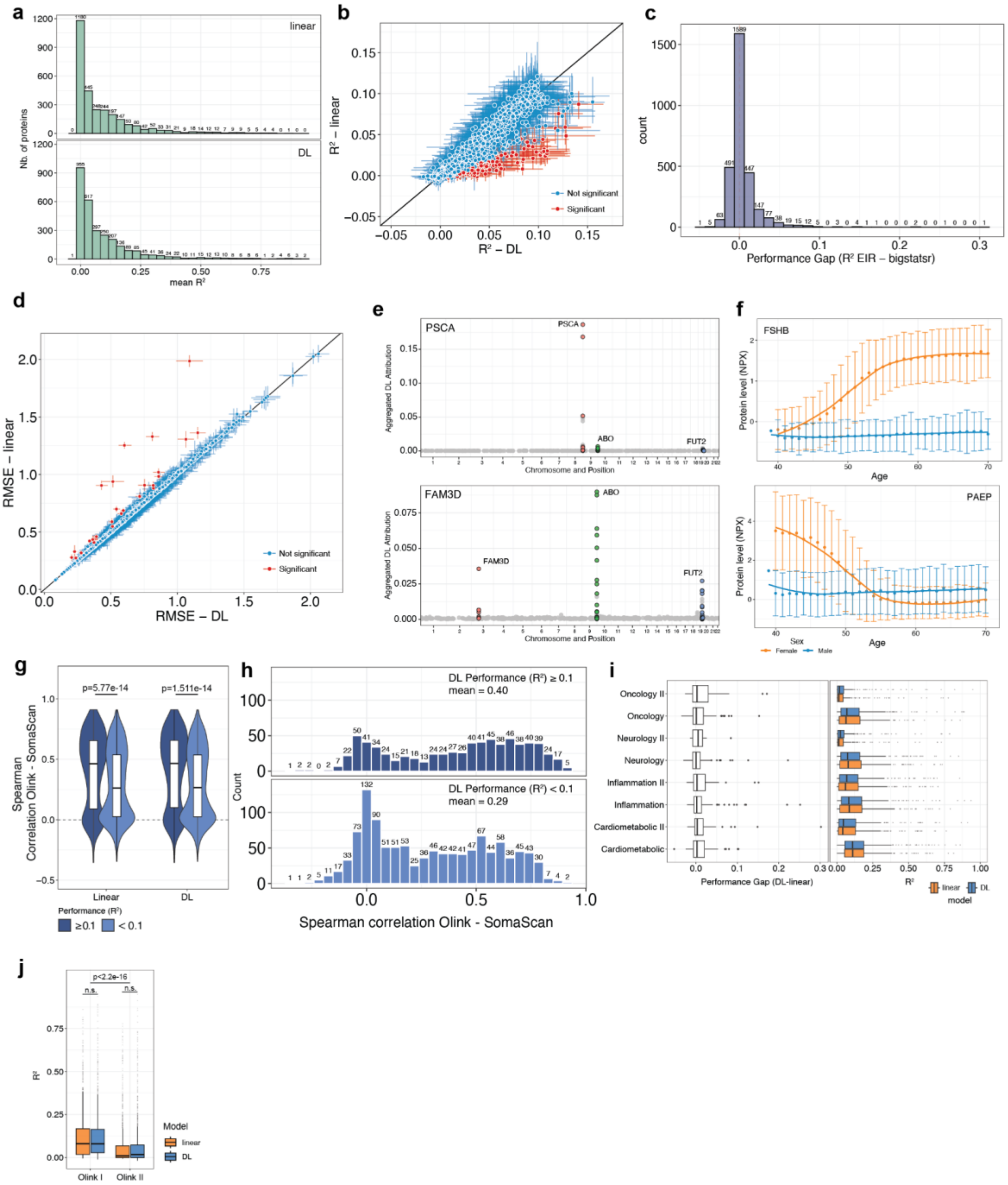


**Supplementary Figure 2**. **a)** Number of proteins and distribution of linear (bigstatsr) and DL (EIR) model performance (mean bootstrapped $R^2$). **b)** DL and linear model performance for proteins with mean bootstrapped $R^2 < 0.1$ in DL or linear model. The error bars indicate the 95% confidence interval from 1000 bootstraps and proteins with non-overlapping confidence intervals between DL and linear models were set as significant and labeled in red. **c)** Number of proteins and their distribution of performance ($R^2$) gap between DL and linear model. Models were trained on n=34,047 individuals and tested on n=1,771 individuals of self-reported UK-white ethnicity. For calculation of performance gap, mean bootstrapped $R^2$ (n=1000 bootstraps) of the linear models were subtracted from the DL models. **d)** DL and linear model performance (Root-mean-squared error (RMSE)) for all 2,922 proteins. The error bars indicate the 95% confidence interval from 1000 bootstraps and proteins with non-overlapping confidence intervals between DL and linear models are called significant and labeled in red. **e)** Aggregated DL attribution of 642 and 757 SNVs across the genome that are used as input to model PSCA (top) and FAM3D (bottom) protein levels. Variants located within the three most activated loci are colored and labeled. **f)** Mean protein expression level (NPX) and Standard deviation across different age groups (n=52,700) in the UKB stratified by sex (male, female) for FSHB (top) and PAEP (bottom). **g)** Spearman correlation between SomaScan and Olink assay of 1,861 proteins from Eldjarn et al. grouped by proteins with high ($R^2 \geq 0.1$) or low ($R^2 < 0.1$) performance in our DL or linear models. Statistical significance was calculated using the Wilcoxon rank-sum test. **h)** Histogram showing the distribution of proteins across Spearman correlation coefficient between SomaScan and Olink. The top panel shows proteins with high ($R^2 \geq 0.1$) DL performance and the bottom panel shows proteins with low DL performance ($R^2 < 0.1$). **i)** Performance gap between DL and linear models (left) and the model performance (mean bootstrapped $R^2$) of DL and linear model (right) of proteins in 8 different panels of the Olink Explore assay. **j)** Model performance (mean bootstrapped $R^2$) of proteins in Olink panel I (Oncology I, Neurology I, Inflammation I, and Cardiometabolic I) or Panel II (Oncology II, Neurology II, Inflammation II, and Cardiometabolic II) in the DL or linear models. Statistical significance was calculated using a two-sided t-test.

### Supplementary Figure 3

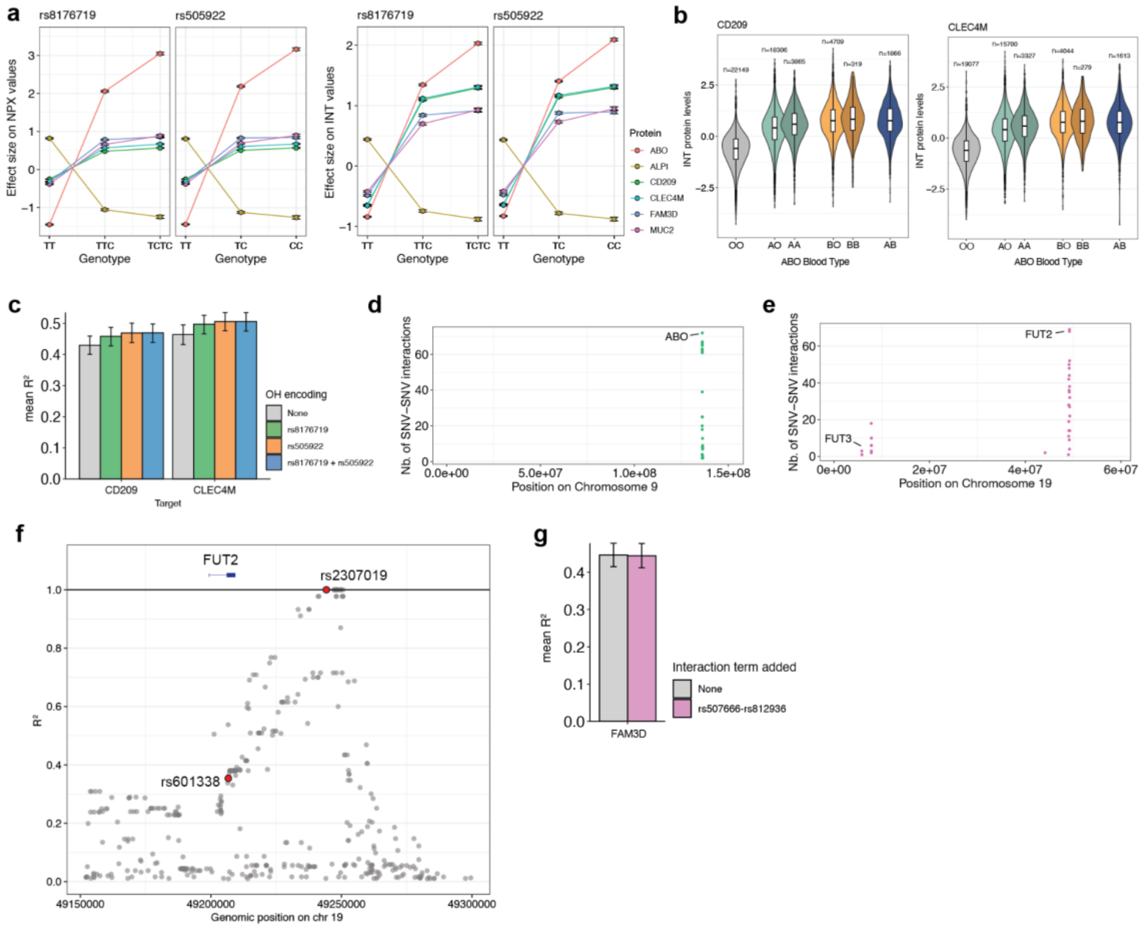


**Supplementary Figure 3. a)** Effect size of different genotypes of *ABO* variants rs8176719 and rs505922 on INT protein levels of ABO, ALPI, MUC2, FAM3D, CD209 and CLEC4M. Error Bars indicate 95% confidence interval. **b)** INT protein levels of CD209 (n=51,214) in individuals in the UK Biobank, stratified by their imputed ABO blood group (field p23165)[54–57]. **c)** Linear model performance to predict CD209 and CLEC4M plasma levels trained on genotypes and covariates. One-hot encoded genotypes rs8176719 and/or rs505922 were added as single terms to assess performance improvement. Error bars indicate 95% confidence intervals of 1000 bootstraps. **d)** Number of interactions per unique SNV for variants on chromosome 9. Location of ABO locus is indicated. **e)** Number of interactions per unique SNV for variants on chromosome 19. Locations of FUT2 and FUT3 loci are indicated. **f)** Proxy variants and their $R^2$ are shown for variant rs2307019 (labeled) based on LDProxy[3,20] calculated for the British population (GBR). FUT2 rs601338 (Trp154Ter) that determines the FUT2 secretor status is labeled and the position of the FUT2 gene (NM_000511) is indicated above the plot. **g)** Linear model performance to predict FAM3D plasma levels trained on one-hot encoded genotypes and covariates. Interaction between rs507666 and rs812936 was added as a single term to assess performance improvement. Error bars indicate 95% confidence intervals of 1000 bootstraps.

### Supplementary Figure 4

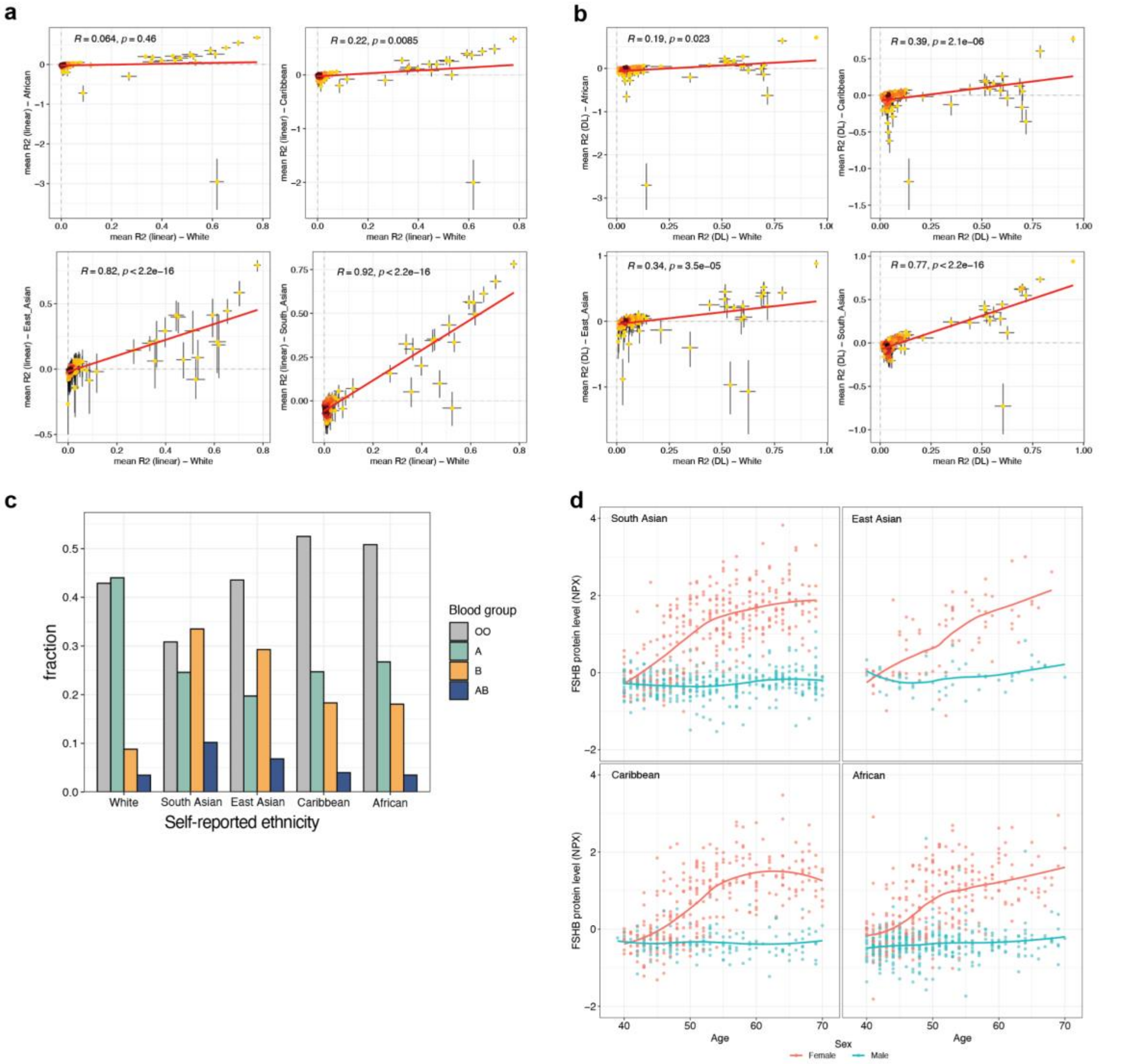


**Supplementary Figure 4. a)** Correlation of mean bootstrapped linear model performance ($R^2$) between test sets of White and African, Caribbean, East Asian or South Asian self-reported ethnicity. Results of pearson correlation test are shown. Error bars indicate 95% confidence intervals of 1000 bootstraps. **b)** Correlation of mean bootstrapped DL model performance ($R^2$) between test sets of White and African, Caribbean, East Asian or South Asian self-reported ethnicity. Results of pearson correlation test is shown. Error bars indicate 95% confidence intervals of 1000 bootstraps. **c)** Distribution of imputed ABO blood groups within the self-reported ethnicity groups in the UKB-PPP. **d)** Age-dependent protein levels for FSHB between female and male individuals of South Asian, East Asian, Caribbean or African self-reported ethnicity test sets.

### Supplementary Figure 5

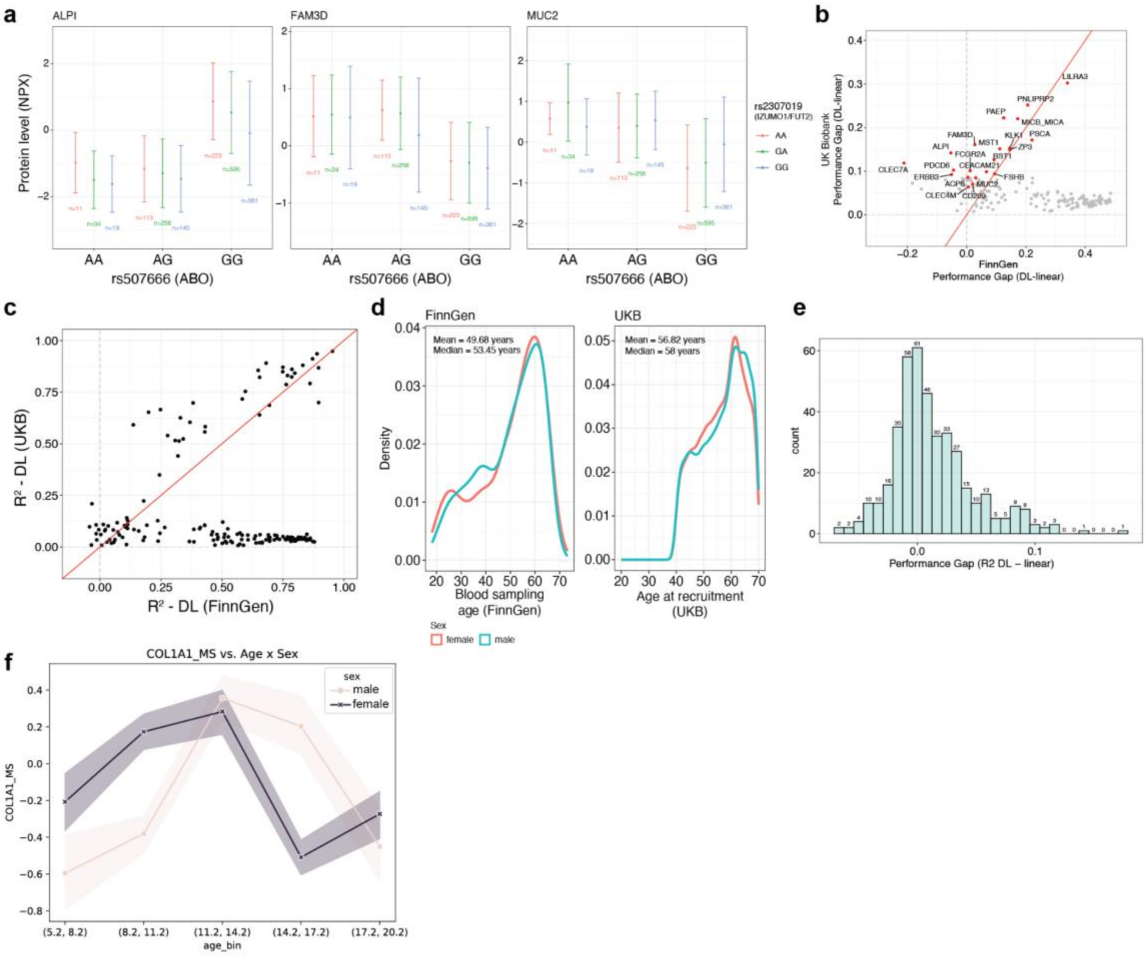


**Supplementary Figure 5. a)** Plasma levels (NPX) of ALPI, FAM3D and MUC2 in individuals from the FinnGen project in all combinations of genotypes of the *ABO* variant rs507666 and the *IZUMO1* variant rs2307019. Error bars indicate standard deviation. Numbers of individuals in the different genotype combinations are indicated. **b)** Correlation of Performance gap (R2-R2) between UKB and FinnGen for 170 proteins. The top 20 proteins based on the absolute performance gap in the UKB as well as CD209 and CLEC4M are labeled and colored in red (**Figure 2d**). **c)** Correlation of DL performance between UKB (mean bootstrapped R2) and FinnGen (mean R2) for 170 proteins. **d)** Distribution of the age at blood sampling stratified by sex in the FinnGen project (left) and distribution of the age at recruitment in 48,594 individuals in the UKB-PPP stratified by sex (right). Mean and median age of all participants in the respective cohort are indicated. **e)** Number of proteins and their distribution of performance ($R^2$) gap between DL (EIR) and linear (bigstatsr) model trained on 1,533 individuals and tested on 190 individuals from the Holbaek study. **f)** Plasma levels of COL1A1 in different age groups between 5.2 and 20.2 years stratified by sex.
